# Public health impacts of an imminent Red Sea oil spill

**DOI:** 10.1101/2021.04.08.21255129

**Authors:** Benjamin Q Huynh, Laura H Kwong, Mathew V Kiang, Elizabeth T Chin, Amir M Mohareb, Aisha O Jumaan, Sanjay Basu, Pascal Geldsetzer, Fatima M Karaki, David H Rehkopf

## Abstract

The possibility of a massive oil spill in the Red Sea is increasingly likely. The *Safer*, a deteriorating oil tanker containing 1.1 million barrels of oil, has been deserted near the coast of Yemen since 2015, and threatens environmental catastrophe to a country presently in a humanitarian crisis. Here, we model the immediate public health impacts of a simulated spill. We estimate all of Yemen’s imported fuel through its key Red Sea ports would be disrupted, and that the anticipated spill could disrupt clean water supply equivalent to the daily use of 9 to 9.9 million people, food supply for 5.7 to 8.4 million people, and 93% to 100% of Yemen’s Red Sea fisheries. We also estimate an increased risk of cardiovascular hospitalization from pollution ranging from 5.8% to 42.0% over the duration of the spill. The spill and its potentially disastrous impacts remain entirely preventable through offloading the oil. Our results stress the need for urgent action to avert this looming disaster.

## Main

Since 2015, war and blockade in Yemen has made it the site of “the world’s worst humanitarian disaster.”^1^ A major consequence has been the abandonment of the *Safer*, a deteriorating oil tanker moored 4.8 nautical miles off the Red Sea coast of Yemen. Concerns of a massive spill have arisen as the *Safer*, designated out-of-class since 2016 and not maintained since the start of the conflict, continues to deteriorate. The *Safer* contains 1.1 million barrels of oil, more than four times the amount spilled in 1989 by the *Exxon Valdez*.^2^ The prospective spill threatens to harm the environment, economy, and public health of the countries bordering the Red Sea.

The possibility of a spill is increasingly likely. The visibly dilapidated *Safer* is single-hulled, meaning a breach will cause the onboard oil to spill directly into the sea. Water entered the engine room in May 2020 through a seawater-pipe leak, and the vessel’s fire extinguishing system is non-operational.^3^ A spill could occur due to a leak or combustion. A leak could arise through continued deterioration of the vessel’s hull, or by breach of the hull due to inclement weather; combustion could occur through build-up of volatile gases aboard the vessel or direct attack on the vessel. Ansar-Allah (colloquially known as the Houthis), a political and armed movement in control of North Yemen, currently has access to the *Safer*. As of writing, negotiations between the United Nations and the Houthis to inspect and repair the *Safer* have stalled indefinitely, and no long-term solutions, such as offloading the oil, have been publicly proposed.

Yemen is particularly vulnerable to the anticipated spill due to reliance on major ports near the *Safer*, Hudaydah and Salif, through which 68% of humanitarian aid enters the country. In the event of port disruption, rerouting humanitarian aid would be logistically difficult due to regional instability, lack of capacity at other ports, and the ongoing blockade, which severely limits the entry of supplies.^4^ Overall, Yemen imports 90-97% of its fuel and 90% of its food supply.^5^ Over half of Yemen’s population is dependent on the humanitarian aid delivered at ports, with 18 million people requiring clean water assistance and 16 million requiring food assistance.^1^

The anticipated spill also threatens the clean water supply of the water-scarce Red Sea region. Oil could contaminate the desalination plants that are lined along the coast north of the *Safer*, thereby disrupting clean water supply to the region at large. For Yemen in particular, clean water is mostly supplied through groundwater pumps or water trucks, both of which require fuel. Previous fuel shortages caused by the blockade resulted in far-reaching public health impacts: for example, clean water and sewage systems stopped operating, solid waste collection was stalled, and electrical grid disruptions led to blackouts affecting hospital operations, all of which contributed to a massive cholera outbreak in 2017.^6^

Yemen’s fisheries, responsible for providing subsistence for 1.7 million people in the country, would also be threatened. Fishing was Yemen’s second largest export before conflict began and continues to provide a source of income and food security in a country on the brink of famine. The sector has substantially declined in recent years due to conflict and fuel shortages; a massive oil spill would devastate an industry already struggling to subsist.^7–9^

Pollution from the spill, whether by evaporation or smoke from combustion, can cause cardiovascular and respiratory health issues. Fine particulate matter (PM_2.5_) in general is known to increase the risk of hospitalization from cardiovascular and respiratory diseases, and pollution from oil spills in particular is known to cause a variety of health issues, ranging from psychiatric to respiratory symptoms.^10–14^ Resultant pollution from a spill could increase the burden on Yemen’s under-resourced health systems and hinder cleanup efforts.

Major oil spills are known to have wide-ranging environmental and economic consequences. The danger that the *Safer* poses to the Red Sea’s unique ecosystem has been documented.^2,15^ However, the immediate public health impacts of the *Safer* spilling remain uncharacterized, and the extent to which the anticipated spill could disrupt humanitarian aid to Yemen critically informs the question of whether and with what urgency interventions should be deployed. Here we model the immediate public health impacts that would follow a major spill from the *Safer*. We stochastically simulate oil spills using historical data to assess likely spill and pollution trajectories, and use these results to estimate disrupted access to fuel, food, and clean water, as well as initial air pollution-related health effects.

### Modeled spills consistently reach key Red Sea ports

We simulate the *Safer* spilling over a variety of historical weather conditions and find most simulated spills tend to move towards Yemen’s northwest coast (Figure 1). We observe seasonal variation from our models: in the summer, spills tend to move southeast and further along Yemen’s coastline, but in the winter, spills tend to move north along the Red Sea coast. Uncertainty in the estimates indicate a wide range of possible trajectories across the Red Sea, showing the possibility of movement in either direction for both seasons. We estimate it will take 6-10 days for the oil to reach Yemen’s western coastline. We estimate 51% (95% uncertainty interval: 46-54%) of the oil will evaporate within 24 hours of leaking from the vessel, with the heavier components remaining on the water (Figure 2). Modeled cleanup attempts -- skimming, *in situ* burns, and dispersants -- remove a negligible amount of oil within the first six days. Under optimistic conditions, cleanup interventions initially reduce evaporation rates slightly (likely due to the delayed impact of dispersants), and by six days we estimate 39.7% of the oil to remain floating, compared to 38.2% under evaporation alone (Figure 2). If the spill spreads unmitigated for three weeks, oil will likely impede passage through the Gulf of Aden (Figure 1).

**Figure 1:**
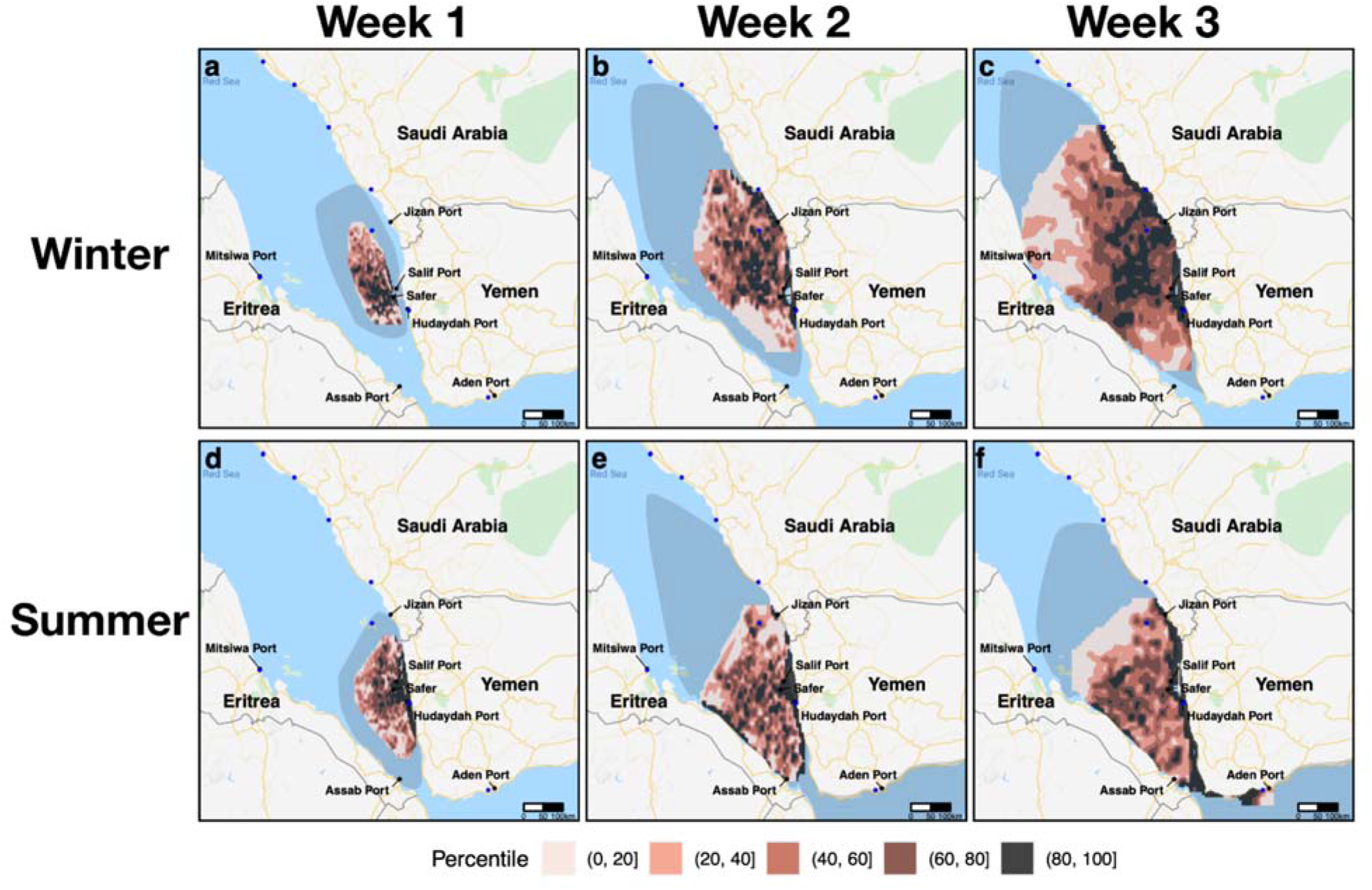
Average surface oil concentration of 1,000 simulated spills in the winter (a,b,c) and 1,000 spills in the summer (d,e,f). Columns denote progress of the 1,000 spills after one week (a,d), two weeks (b,e), and three weeks (c,f). Colored contours represent percentiles of average surface concentration over 1,000 simulated spills, and can be interpreted as the expected surface concentration relative to other grid cells in the exposed area. Shaded region represents the area within which approximately 90% of spill trajectories are expected to fall. Blue dots represent desalination plants.

**Figure 2:**
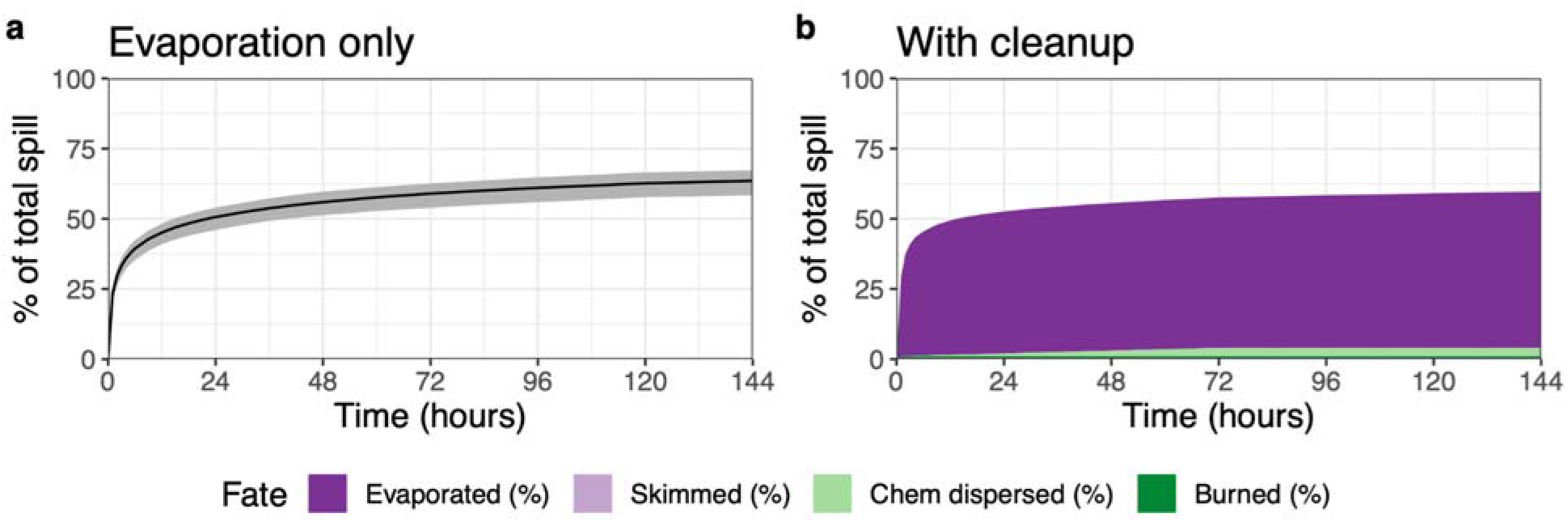
Oil fate scenarios. (a) Oil spill evaporation over time. Black line indicates mean, and gray ribbon indicates 95% uncertainty interval, defined as the 2.5^th^ and 97.5^th^ percentiles of values across 1,000 Monte Carlo simulations. (b) Oil fate under optimistic cleanup conditions. Colored regions denote different oil removal methods.

Ports and desalination plants, crucial for providing fuel, food, and water, stand to be disrupted by the spill. We estimate that two weeks after a spill Yemen’s key ports of Hudaydah and Salif will likely be directly impacted, with average surface oil concentrations that are in the 90th percentile compared to other exposed areas (Supplementary Table 1). We estimate that by three weeks, a spill could even reach as far as the port of Aden (outside of the Red Sea), and the desalination plants and ports in Eritrea and Saudi Arabia (Figure 1, Supplementary Table 1, Supplementary Figures 1-6).

### Expected port closures disrupt access to aid

The spill and subsequent port closures will disrupt maritime transport across the Red Sea, rerouting many shipments around Africa. We estimate that for each month of Red Sea port closure, delivery of 200,000 (180,000-250,000) metric tons of fuel for Yemen will be disrupted, equivalent to 38% of the national fuel requirement.^16^ We consequently expect fuel prices in Yemen to spike; when the blockade was tightened to fully close ports in November 2017, fuel prices sharply rose across the country, with prices in Hudaydah rising by 72% in the following month (Figure 3).

**Figure 3:**
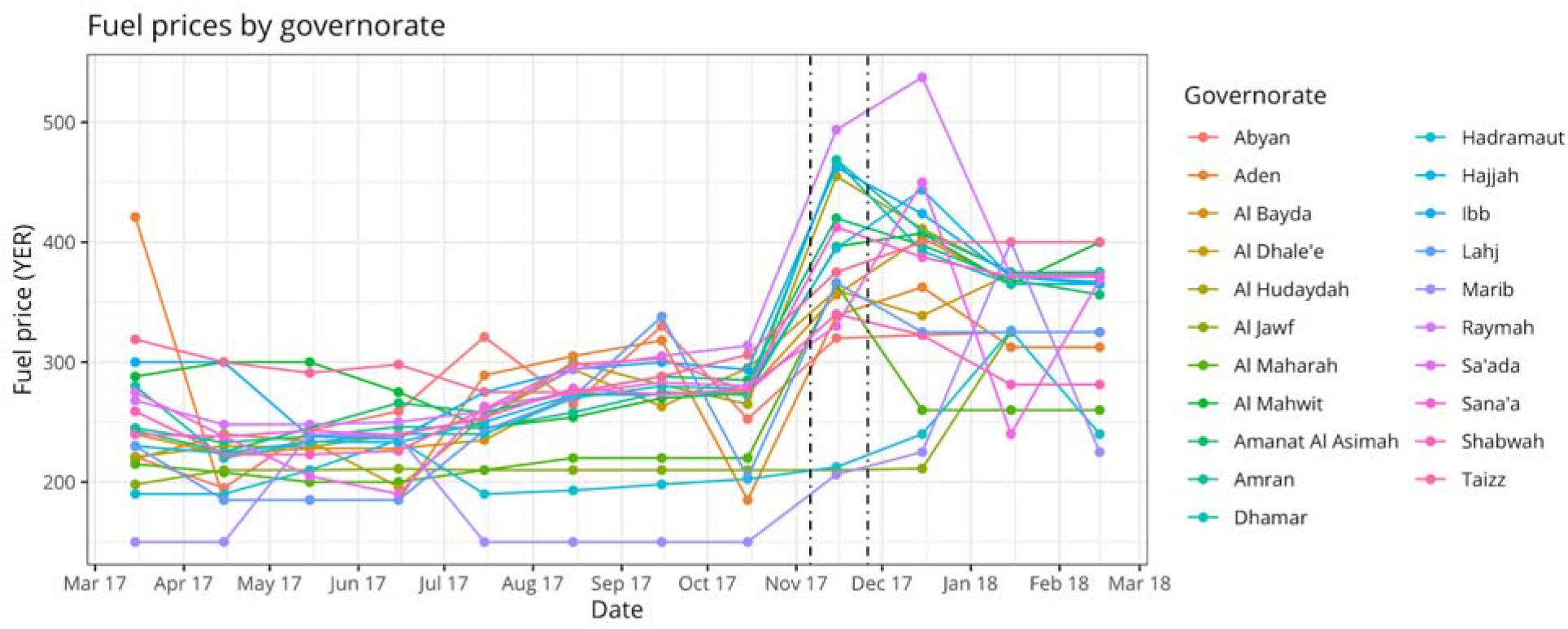
Fuel prices by governorate, March 2017 - March 2018. Dashed lines denote the start and end of full port closures in Yemen.

The oil spill will also threaten clean water supply, equivalent to the daily use of an estimated 1 to 1.9 million people through potential contamination of desalination plants. In the Red Sea region, we estimate potential disruption of desalination plants responsible for a total of 77,000 m^3^/day of clean water in the summer, and 362,000 m^3^/day in the winter (Supplementary Table 2). We additionally expect water access in Yemen to be severely disrupted by fuel shortages if the spill closes ports; during the full port closures in November 2017, 8 million people in Yemen lost access to running water since accessing water typically depends on fuel-powered pumps or water trucks.^5,17^

Similarly, food security will be threatened by potential food aid disruptions and fishery closures. In the event of Yemen’s Red Sea ports closing within two weeks of the spill, food aid will be disrupted for an estimated 5.7 (3.7-8.1) million people who currently require food assistance. We estimate that if Aden’s port also closes, a total of 8.4 (5.4-11.9) million people will not receive food aid. We estimate the spill threatens 66.5% to 85.2% of Yemen’s Red Sea fisheries within one week, and 93% to 100% of Yemen’s Red Sea fisheries within three weeks, depending on season. In the summer, even 2.6% of Yemen’s Gulf of Aden fisheries are threatened within three weeks (Supplementary Table 3).

### Air pollution increases hospitalization risk

We predict moderate short-term health effects from air pollution, with estimates for the average increased risk of cardiovascular and respiratory hospitalizations ranging from 5.8% (0-7.5) across 11.3 million (0-27) person-days for a slow-release winter spill to 31.2% (6.5-50.5) across 19.5 million (0.4-24.2) person-days for a fast-release summer spill (Table 1). Combustion would increase pollution, with estimates for the average increased risk of cardiovascular and respiratory hospitalizations ranging from 6.7% (5.2-7.9) across 22.3 million (1.2-41.8) person-days for a slow-release winter spill to 42.0% (21.9-61.4) across 22.7 million (17.0-26.0) person-days for a fast-release summer spill. Seasonality effects are present, with simulated air pollution traveling east into Yemen during the summer, and west into the sea during the winter (Figure 4). In some winter simulations, pollution did not reach the Yemeni population at all. Air pollution is highest in the immediate vicinity of the oil, reaching PM_2.5_ levels as high as 1600 ug/m^3^, corresponding to a 530% (460-590) increased risk of cardiovascular and respiratory hospitalization for individuals directly exposed to the oil, such as cleanup workers.

**Table 1:** Population-weighted average increased risk (IR) and exposed populations for cardiovascular and respiratory hospitalizations from air pollution over various scenarios and spill durations. Spill duration is equivalent to exposure duration. All intervals denote 95% uncertainty intervals, defined as the 2.5^th^ and 97.5^th^ percentiles of simulated values. Exposed population is defined as having been exposed to 10 ug/m^3^ or more of PM_2.5_. Uses estimates from Burnett et al.^53^

| Scenario | Average IR (%) | Person-days |
| --- | --- | --- |
| Summer, Fast-release, Leak | 31.24<br>(6.52,50.56) | 19.46<br>(0.38,24.18) |
| Winter, Fast-release, Leak | 16.2<br>(6.82,22.59) | 4.52 (0.85,8.73) |
| Summer, Slow-release, Leak | 13.9<br>(3.95,22.01) | 53.29<br>(0.11,79.87) |
| Winter, Slow-release, Leak | 5.83 (0,7.48) | 11.34 (0,27.19) |
| Summer, Fast-release, Fire | 41.96<br>(21.88,61.41) | 22.65<br>(17.02,26) |
| Winter, Fast-release, Fire | 19.45<br>(13.15,24.72) | 6.6 (2.39,9.98) |
| Summer, Slow-release, Fire | 17.72<br>(10.37,25.09) | 68.21<br>(36.95,82.63) |
| Winter, Slow-release, Fire | 6.65 (5.16,7.9) | 22.34<br>(1.15,41.75) |

**Figure 4:**
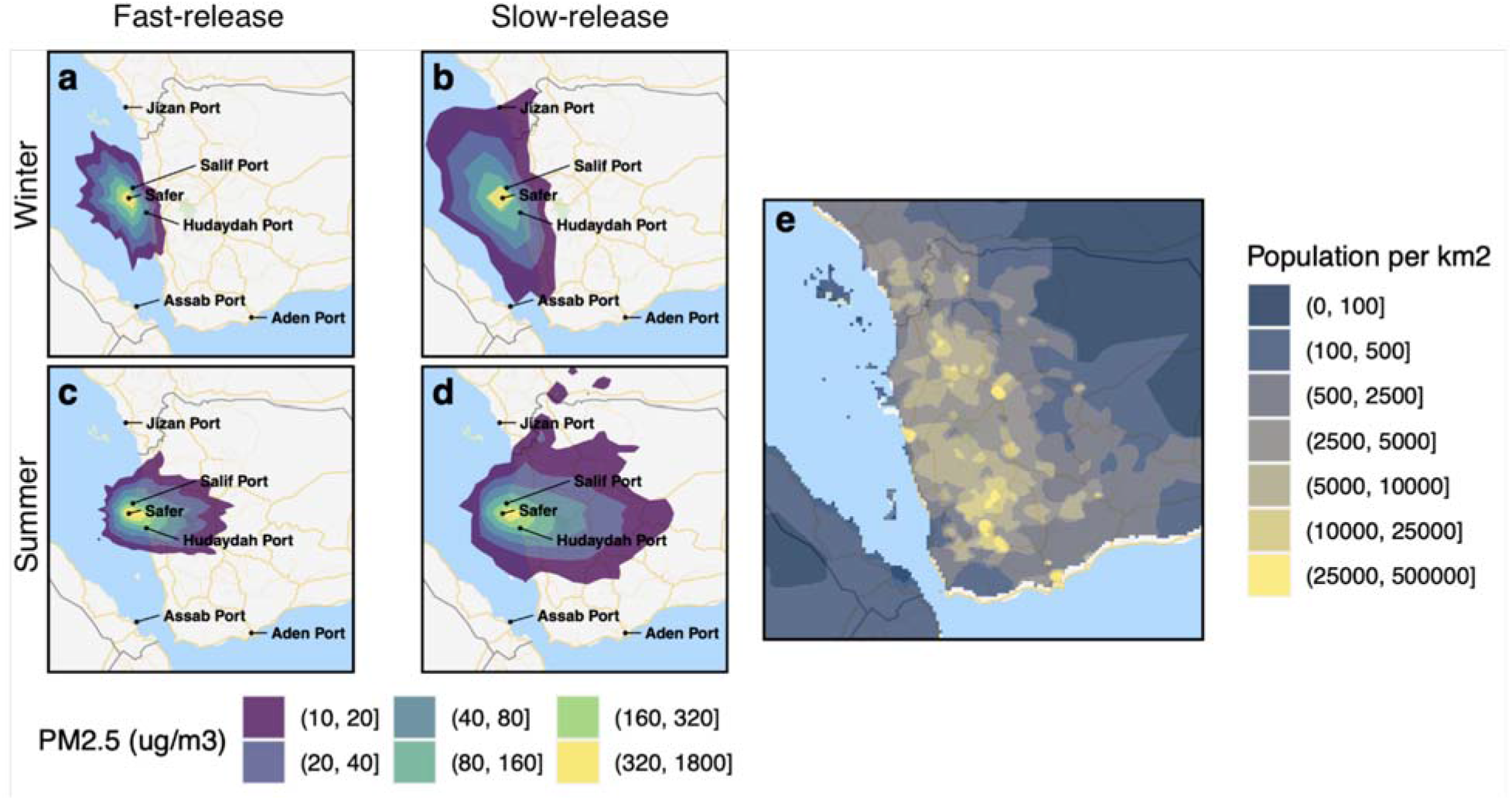
Projected 24-hour average air pollution concentration at the end of spill in the winter (a,b) and summer (c,d) for fast-release spills (a,c) and slow-release spills (b,d), alongside a population density plot (e).

## Discussion

The public health impacts of a spill from the oil tanker *Safer* are expected to be catastrophic, particularly for Yemen. Disruption of fuel imports is anticipated to shut down hospitals and essential services, at a time when Yemen already faces fuel shortages and only 50% of its health services are functional.^18,19^ Both fuel shortages and contamination of desalination plants are expected to worsen an existing water crisis, potentially leading to a resurgence of water-borne infectious diseases.^20,21^ Disruption to food aid would likely increase food prices and exacerbate an ongoing famine.^1^ The spill threatens to disrupt nearly all of Yemen’s Red Sea fisheries, which would worsen food security and exacerbate Yemen’s displacement crisis as workers seek new employment.^7,19^ Imports of medical supplies from aid groups would likely also be disrupted, further destabilizing health services.^8,9^ Our modeling indicated that cleanup would be slow even under optimistic conditions; actual cleanup efforts would potentially be further prolonged and logistically difficult given the conflict in the region and sea mines in the water. Ports would potentially remain closed until sufficient cleanup has occurred; if ports were to prematurely reopen, ships would risk remobilizing the oil and furthering environmental damage. The air pollution from the spill may be moderate in comparison to the supply disruptions from the spill, but clean-up workers, essential to curbing impacts of the spill, may be at high risk of hospitalization,^13,25,26^ and the pollution with its resultant increases in respiratory hospitalizations would potentially further strain an already under-resourced healthcare system.^27^ Personal protective equipment could substantially mitigate pollution harms, but the ongoing medical equipment shortage would likely be exacerbated by port closures.^28,29^

The long-term and global impacts of the spill, although outside the scope of our modeling analysis, are also potentially severe. Ecological and environmental impacts through wildlife endangerment and coastline contamination from large oil spills can persist for years or decades.^56,57^ In particular, the spill threatens the Red Sea coral reefs, studied for their unique resilience to seawater warming.^30^ Further, the spill could hinder global trade through the vital Bab el-Mandeb Strait, 29 kilometers wide at its narrowest point, through which 10% of the global shipping trade passes. Exclusion zones created for cleanup could reroute traffic, and shipments will be delayed as ships potentially exposed to oil will require cleaning.

Kleinhaus et al warned of the danger the *Safer* poses to the Red Sea’s ecosystem, and performed single-simulation analysis, finding that particles in February flow north and particles in August flow south.^2^ Our spill trajectory analysis strengthens their findings by explicitly modeling the *Safer*’s oil properties and thoroughly assessing the range of possible spill trajectories through thousands of Monte Carlo simulations over historical data. We find their simulated trajectories to be consistent with ours, and we validate their claim that winter spills tend to travel north and summer spills tend to travel south. However, we emphasize that our primary finding, that a spill from the *Safer* could lead to catastrophic public health impacts, is true regardless of season due to the wide range of spill trajectories that we observe from our uncertainty analyses. Relatedly, a misinterpretation of seasonality may lead malicious actors to induce an oil spill during supposedly favorable conditions; we assert strongly that our analysis shows that substantial uncertainty in trajectory exists despite seasonality, and so all parties involved in the conflict stand to bear the burden of the spill.

Although our model accounts for a number of uncertainties associated with a future spill, some uncertainties remain. Our oil spill model is averaged over simulations based on historical data, but spills often occur due to extreme conditions, so the actual spill may manifest differently than the scenarios we present. The data we use to model the spill and downstream effects are of variable quality, so our results may be affected by measurement error. Stored quantities of supplies could mitigate shortages in the event of port disruption, but such quantities are likely minimal as Yemen is already undergoing supply shortages, and price spikes would render supplies inaccessible for many.^5,24^ Our model only considers the scenario of a full spill; we do not model, for example, the impacts of a minor leak in the hull. We do not provide expected durations for port and desalination plant closures since we are unable to predict the timing of cleanup efforts. In particular, it is difficult to predict the international response to the anticipated spill: some parties may prefer ports to remain closed to minimize environmental damage, and others may prefer them to open sooner to minimize supply disruptions. Our cleanup analysis does not consider how containment booms may change the spill trajectory, but booms would have limited ability to change the spill trajectory due to the size of the spill, wave heights in the Red Sea, and lack of pre-existing capacity to respond immediately.^31,32^ Our spill models do not explicitly model tides, and instead assumes tidal currents to be implicitly measured in the currents data; the Red Sea has a small tidal range, so we do not expect this to substantially affect our model estimates.^33^ Our plotted uncertainty regions for potential spill trajectories correctly identify the outer uncertainty particles of where spills may go, but may overestimate the areas within the region, as there may be pockets of sea where oil is less likely to travel due to patterns in winds or currents. However, our models also do not fully account for wave turbulence, so the affected spill area may be larger than we show. We also make the conservative assumption that ports will only be disrupted when oil directly reaches them. In reality, oil only needs to impede nearby maritime traffic to threaten port closure, so as to prevent the environmental hazard of ships remobilizing the oil. Although we are unable to predict the exact probability of port closure, our spill analysis validates the claim that port closures are a distinct possibility, as agencies familiar with the situation have warned.^34^ We thus do not believe that uncertainty in our spill models should qualitatively change our assessment of the potential downstream public health impacts as severe.

Our air pollution models are also subject to several levels of uncertainty. Our evaporation and combustion estimates are partially based on data from the *Deepwater Horizon*, which had light crude oil that was similar but not equivalent to the oil carried by the *Safer*. It is possible that the tanks aboard the *Safer* have already leaked, and that some of the light components of the oil have already evaporated, which would reduce the amount of air pollution from the spill. We based our health impact assessments on single-pollutant health outcomes and do not explicitly model volatile organic compounds due to lack of data, but the PM_2.5_ from an oil spill is likely different from PM_2.5_ from other sources. Our estimates of the size of the population affected by pollution may be subject to measurement error in the original population dataset resulting from displacement or regional instability. We also only consider the risk of cardiovascular and respiratory hospitalization from oil exposure; oil spills are known to also have neurological, hematological, dermatological, and psychiatric impacts.^13,14,26^ Our modeling thus does not capture the full extent of health effects from oil exposure. Further, our modeling assumed the pollution is emitted from a single site (i.e. the site of the spill), but in reality it would be emitted from wherever the oil is spilled. Thus, our estimates of hospitalization rates are biased downwards, since the pollution would likely be slightly closer inland.

Despite the uncertainty inherent to our modeling, our evidence demonstrates that an oil spill from the *Safer* poses an extreme public health risk to the people of the surrounding area, with Yemen bearing the largest impact. Our results show the spill will jeopardize access to fuel, food, and water in Yemen, a country already facing shortages of all three. Other countries bordering the Red Sea will also incur the burden of the spill with port closures and disruptions to desalination plants. This public health disaster could be averted by finding a long-term solution to handling the oil aboard the *Safer*, underscoring the need for urgent action from the international community.

## Methods

### Data

For gridded wind data, we use the 2019 and 2020 surface wind ERA5 datasets from the European Centre for Medium-Range Weather Forecasts (ECMWF), with a 1-hour temporal resolution and 1/4° spatial resolution.^35^ For gridded currents, sea temperature, and salinity data, we use 2019 and 2020 data from the Hybrid Coordinate Ocean Model, with a 3-hour temporal resolution and 1/12° spatial resolution.^36^ For data on the properties of the oil, we use NOAA’s Oil Library Project.^37^ For data on fuel prices, we use a dataset from the World Food Programme.^38^ We use fuel import data from the United Nations Verification and Inspection Mechanism for Yemen.^39^ We used a variety of sources for data on desalination plant locations and capacity (see Supplementary Table 2).^40–42^ For Yemen, locations for all desalination plants were not available, and the water capacities for the known plants were also unavailable. Therefore to estimate the water capacity for each of the known plants in Yemen, we used the most recent available data on country-wide desalination capacity,^43^ and divided it equally amongst the known plants. Food import estimates were derived from Yemen’s port data.^44^

### Oil spill modeling

We modeled the spill using the pyGNOME library from the National Oceanic and Atmospheric Administration (NOAA) to use their GNOME model.^45^ GNOME is a widely-used Eulerian/Lagrangian spill-trajectory model, modeling spills with Lagrangian elements within flow fields. Like most operational response tools, GNOME is able to model the oil transport and weathering processes of advection, diffusion, dispersion-entrainment, emulsification, evaporation, spreading, oil-shoreline interaction, and dissolution. We chose NOAA’s suite of modeling tools due to their history of being implemented operationally and validated against real-life environmental disasters as well as their widespread use among disaster response agencies.^46^ As model inputs, we used the characteristics of the crude oil onboard the *Safer*, Marib Light, as well as the historical currents and wind data of the region. We performed 1,000 Monte Carlo simulations each for both summer (June-August) and winter (December-February), varying time of day and date. Seasons were chosen based on known current patterns in the Red Sea.^47^ We restricted simulations to three-week timelines due to predictability limits inherent to oil spill modeling and uncertainty in clean-up efforts.^48^

Each simulation had 1,000 particles representing the oil trajectory based on historical weather conditions. Particles were sizeless and represented by surface oil concentration values and coordinates on a latitude-longitude grid rounded to three decimals, approximately corresponding to 100 meters by 100 meters. For each season, we simulated 1,000 spills with three-week timelines. At the end of weeks 1, 2, and 3, we calculated the average surface concentration value at each point on the grid. We then used bilinear interpolation to calculate values between points. Given the uncertainty in the amount of oil that will be spilled, we converted the average surface concentration values from absolute values (average surface oil thickness, measured in meters) to relative values (average surface oil thickness relative to other exposed areas, measured in percentiles).

Each simulation also had 1,000 uncertainty particles that simulated the oil trajectory through parameter settings that assume extreme weather conditions. According to GNOME documentation, the area enclosed by the uncertainty particles represents where approximately 90% of spill trajectories are expected to fall.^49^ We calculate the area enclosed by the uncertainty particles by computing the convex hull from the locations of all uncertainty particles from all 1,000 simulations, and plotted it as the uncertainty region.

We repeated our spill analyses, varying spill duration, season, grid resolution, and number of particles to assess how model output would change. Our original models assumed a 7-day spill; we repeated the models for a 24-hour spill. We further repeated the spill models for spring (March-May) and autumn (September-November). We also resimulated the spills using a coarser spatial resolution (latitude and longitude rounded to 2 decimal points, corresponding to approximately 1.1 by 1.1 kilometers) and higher number of particles (10,000 instead of 1,000). See Supplementary Information.

### Oil Fate

To calculate the fate of the oil, we used NOAA’s Automated Data Inquiry for Oil Spills tool. As inputs, we used data on the oil type, gridded winds, gridded currents, water salinity, and water temperature. We modeled oil fate for two scenarios: one with no cleanup (evaporation only), and one with extremely optimistic cleanup conditions. We restricted all weathering models to run for six days because they do not account for longer-term factors, such as biodegradation or photooxidation, that may affect the weathering rate.^45^ To get a range of evaporation estimates, we performed 1,000 Monte Carlo simulations by randomly varying time and date. We calculated the mean and 95% uncertainty intervals, defined as the 2.5th and 97.5th percentiles across all simulations. For clean-up analyses, we made the optimistic assumptions that (1) cleanup would begin immediately after the spill occurs, and (2) oil recovery amounts over six days would be comparable to those of the 2010 *Deepwater Horizon* oil spill cleanup. We modeled a skimmer with a recovery rate of 14 barrels per hour and 100% efficiency, *in situ* burning with an area of 70,000 m^2^ and 50% efficiency, and 15% of oil sprayed with dispersant at 20% efficiency, at 29 degrees Celsius and 42% salinity. The clean-up parameters were chosen to match the cleanup rate in the *Deepwater Horizon* oil spill. We used the amount of oil recovered by different cleanup methods in the *Deepwater Horizon* oil spill, scaled it from its 85-day cleanup timeframe to our 6-day timeframe, and selected cleanup parameters that would recover this estimated amount of oil over 6 days.^50^ The weather values were selected to match the values from our prior Monte Carlo simulation analysis that maximized oil evaporation.

### Air pollution modeling

For air pollution modeling, we used NOAA’s Hybrid Single-Particle Lagrangian Integrated Trajectory (HYSPLIT) model.^51^ HYSPLIT is an extensively used atmospheric transport and dispersion modeling framework, using a hybrid Eulerian/Lagrangian approach to compute the trajectory of airborne particles as well as pollutant concentrations. We conducted simulations across four scenarios: varying seasons (summer and winter) and spill duration (24-hours for a fast-release spill and 72-hours for a slow-release spill). The spill duration is equivalent to the duration of pollutants being emitted. We used HYSPLIT’s daily runs feature to conduct simulations for every 3 hours from January to March and June to August. Each simulation ran for 144 hours with pollutants emitted at sea level. Each simulation had 2,500 particles, with a concentration value for each particle. For each season and spill duration combination, we calculated the average pollutant concentration value at each point on a latitude-longitude grid rounded to three decimals. We used bilinear interpolation to calculate values between points. We assumed a full spill of 150,000 metric tons in all scenarios.

To get air pollution values in terms of fine particulate matter, we converted the initial oil release from barrels to micrograms, multiplied by the oil to particulate matter conversion rate calculated by Middlebrook et al.,^52^ divided by the duration of the spill, and multiplied by the concentration values estimated by the model. We calculated the population-weighted average increased risk of cardiovascular and respiratory hospitalization from the air pollution by multiplying the air pollution values by the increased risk and population share at each interpolated grid point. Our risk function relating PM_2.5_ exposure to cardiovascular and respiratory hospitalizations was derived from Burnett et al.^53^ Risk was averaged over person-days to compare across different spill durations. We calculated person-days by calculating the number of people exposed to at least 10 ug/m^3^ of PM_2.5_ and the duration for which they were exposed.

Due to uncertainty in the particulate matter conversion rate and increased risk of cardiovascular and respiratory mortality from fine particulate matter, we repeated this process through Monte Carlo simulation, varying those two parameters over 1,000 simulations each to propagate uncertainty. To propagate uncertainty, we varied two parameters over the simulations: the rate of conversion from surfaced oil to fine particulate matter, and the increased risk (IR) of fine particulate matter with respect to cardiovascular and respiratory hospitalizations. Based on the point estimates and confidence intervals (CIs) reported from existing literature, we adopted the methodology from Khomenko et al^54^ and computed the IR standard deviations as:

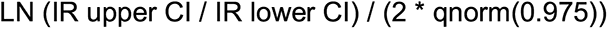

We assumed normal distributions and sampled from the reported mean and estimated standard deviation. The standard deviation for the fine particulate matter conversion rate is provided so we used the reported estimate. We calculated the mean IR from our simulations and constructed 95% uncertainty intervals. If the lower uncertainty interval for the simulated IR was less than 0, we assumed no increased risk of hospitalization. We also calculated the mean and 95% uncertainty intervals for the population affected over time, measured in person-days.

We repeated our air pollution analyses under several different scenarios. In addition to modeling 24-hour spills and 72-hour spills during different seasons, we also modeled air pollution with and without combustion. For estimates of air pollution from combustion, we added the burned oil to particulate matter conversion rate from Middlebrook et al to the existing evaporative conversion rate. We also varied the increased risk function, since the estimate we used from Burnett et al may not demographically reflect the population in Yemen. We thus performed the above calculations using respiratory hospitalization rates from Wei et al and short-term PM_2.5_-related mortality rates from Kloog et al.^10,11^ Wei et al use a Medicare population (mostly aged 65+) that is more vulnerable to air pollution -- which may reflect some subgroups in the Yemeni population, many of whom are malnourished and lack proper health services. Kloog et al use full mortality records from the state of Massachusetts, use more modern methods than Burnett et al, and may reflect the younger age-structure of Yemen than the Medicare population studied by Wei et al. We present results in terms of increased risk of hospitalizations instead of increased hospitalizations, because the increased risk we calculate is relative to a baseline level of hospitalization risk, which we do not know for the population of Yemen. (Supplementary Tables 4 and 5)

### Supply disruption

We estimated fuel disruption by calculating the average and 95% uncertainty intervals of monthly fuel imports through Red Sea ports from January to May 2020 (n = 5), prior to fuel imports being restricted. We calculated the fuel price increase from the November 2017 port closures in Al Hudaydah by taking the median price among diesel, petrol, and gas and comparing it between October 15 2017 and November 15 2017 price data.

We estimated disruption to desalination capacity by compiling a dataset of locations and water capacity of all known plants in the region and identifying locations reached by the simulated oil spills (Supplementary Table 1). Total water consumption equivalents were computed by multiplying each affected country’s share of water amounts by their respective per capita daily usage.

We estimated average and 95% uncertainty intervals of food disruption based on historical data of imports at Yemeni ports.To calculate the amount of food aid disruption, we used 2019 data showing percentages of total food aid in Yemen originating from Hudaydah and Aden.^44^ We then multiplied these percentages by the average of total people targeted for food assistance based on available situation reports from the World Food Programme ranging from March 2020 - February 2021 (n = 10). We used linear interpolation through the *quantile* algorithm in R to construct the uncertainty intervals. See Supplementary Table 6 for estimates as reported and as originally calculated.

We estimated fish yield loss based on gridded annual fish yield in the Red Sea and Gulf of Aden from 2016.^55^ We first filtered for fish yield for only Yemen as a fishing entity, then summed over all types of fishing sectors (artisanal, industrial, etc.). We then summed Yemen’s fish yield over each gridded cell reached by the simulated spill at each week and season. By default, we only included cells if the oil concentration in them was in the 10th percentile of surface concentration or higher so as to exclude cells with trace amounts of oil. To assess how threshold values would affect fish yield loss estimates, we repeated the analysis at no threshold and a threshold of the 20th percentile (see Supplementary Information).

### Data availability

The raw data used during this study are publicly available and described in the supplementary information section. The simulated data are available at https://doi.org/10.7910/DVN/XPESLB.

### Code availability

The code used to support this study is available from the corresponding author upon reasonable request.

**Correspondence and requests for materials** should be addressed to BQH.

## Data Availability

All data used are publicly available.

## Acknowledgments

BQH acknowledges support by the National Science Foundation Graduate Research Fellowship under Grant No. DGE 1656518 and the National Library of Medicine under Training Grant T15 LM 007033. ETC acknowledges support by the National Science Foundation Graduate Research Fellowship under Grant No. DGE 1656518. AMM received funding from National Institutes of Health (NIH) NIAID T32AI007433. PG was supported by the National Center for Advancing Translational Sciences of the NIH under Award Number KL2TR003143. The contents of this article are solely the responsibility of the authors and do not necessarily represent the official views of the NIH. Funding sources had no role in the writing of this manuscript or the decision to submit for publication.

## Author contributions

BQH conceived the initial study design, performed the analysis, interpreted the results, and wrote the initial draft. LHK, MVK, ETC, SB, PG, and DHR helped revise the study design and interpretation of results. AMM, AOJ, and FMK provided crucial contextual information. All authors contributed significantly to writing the final version of the manuscript.

## Supplementary Information

### Supplementary Analyses

Here we present additional sub-analyses showing how our estimates change when varying certain input parameters. Supplementary Figures 1, 2, 3, 4, and 5 are qualitatively similar to Figure 1, showing that varying spill duration (from 7 days to 24 hours), season (from winter and summer to autumn and spring), spatial resolution of spill particles (from 3 rounded decimal points of a longitude-latitude grid to 2), and number of spill particles (from 1000 to 10,000) do not substantially change our results in terms of simulated spills.

Supplementary Table 3 shows how varying the threshold of oil exposure moderately varies the amount of fisheries threatened by the oil spill. Supplementary Tables 4 shows increased hospitalization risk from air pollution using a different risk factor calculation, and Supplementary Table 5 shows increased mortality risk from air pollution.

**Supplementary Figure 1:**
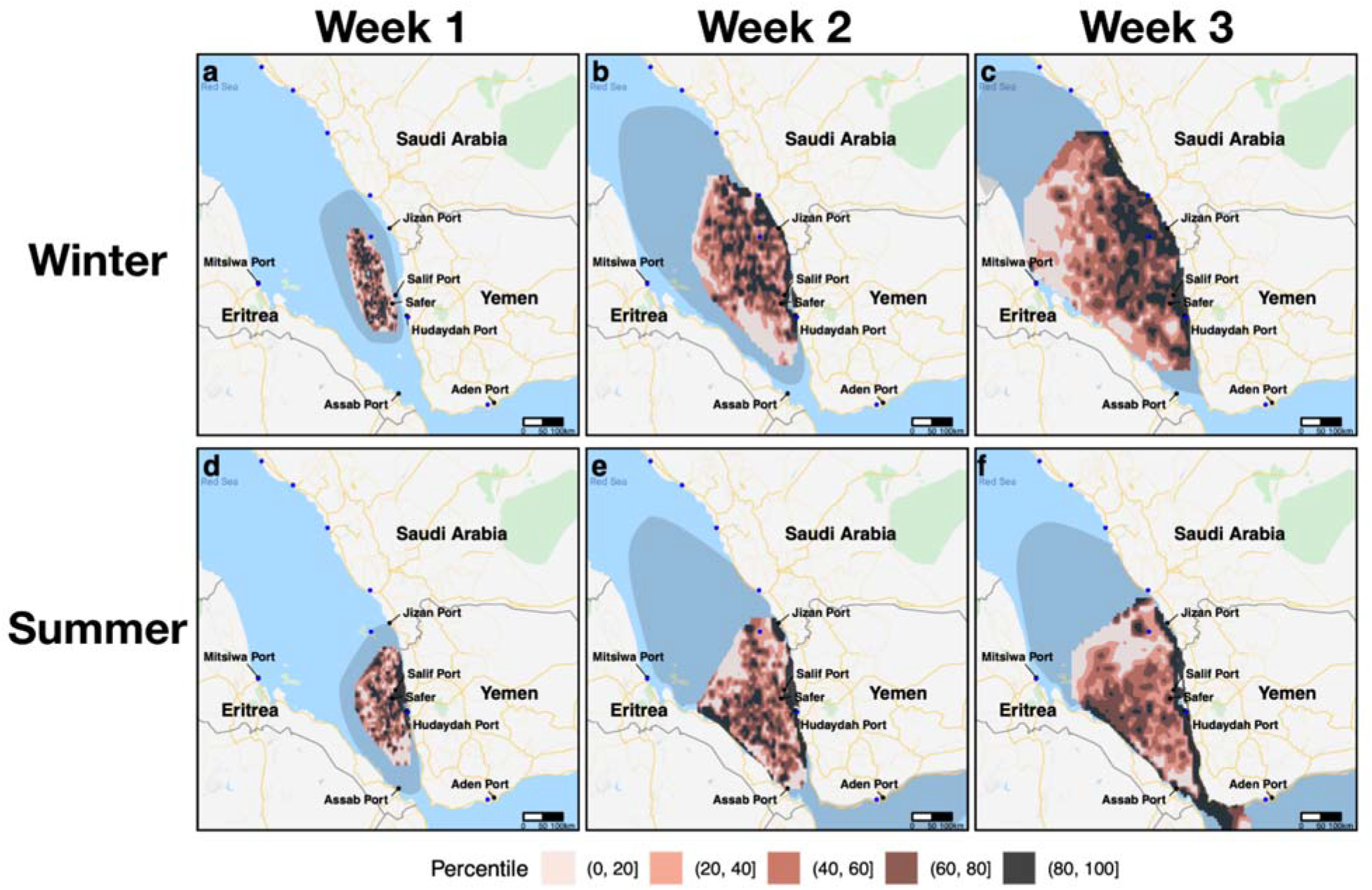
Average surface oil concentration of 1000 simulated spills in the winter (a,b,c) and 1000 spills in the summer (d,e,f), assuming a 24-hour spill duration. Columns denote progress of the 1000 spills after one week (a,d), two weeks (b,e), and three weeks (c,f). Colored contours represent percentiles of average surface concentration over 1000 simulated spills. Shaded region represents the area within which approximately 90% of spill trajectories are expected to fall. Blue dots represent desalination plants.

**Supplementary Figure 2:**
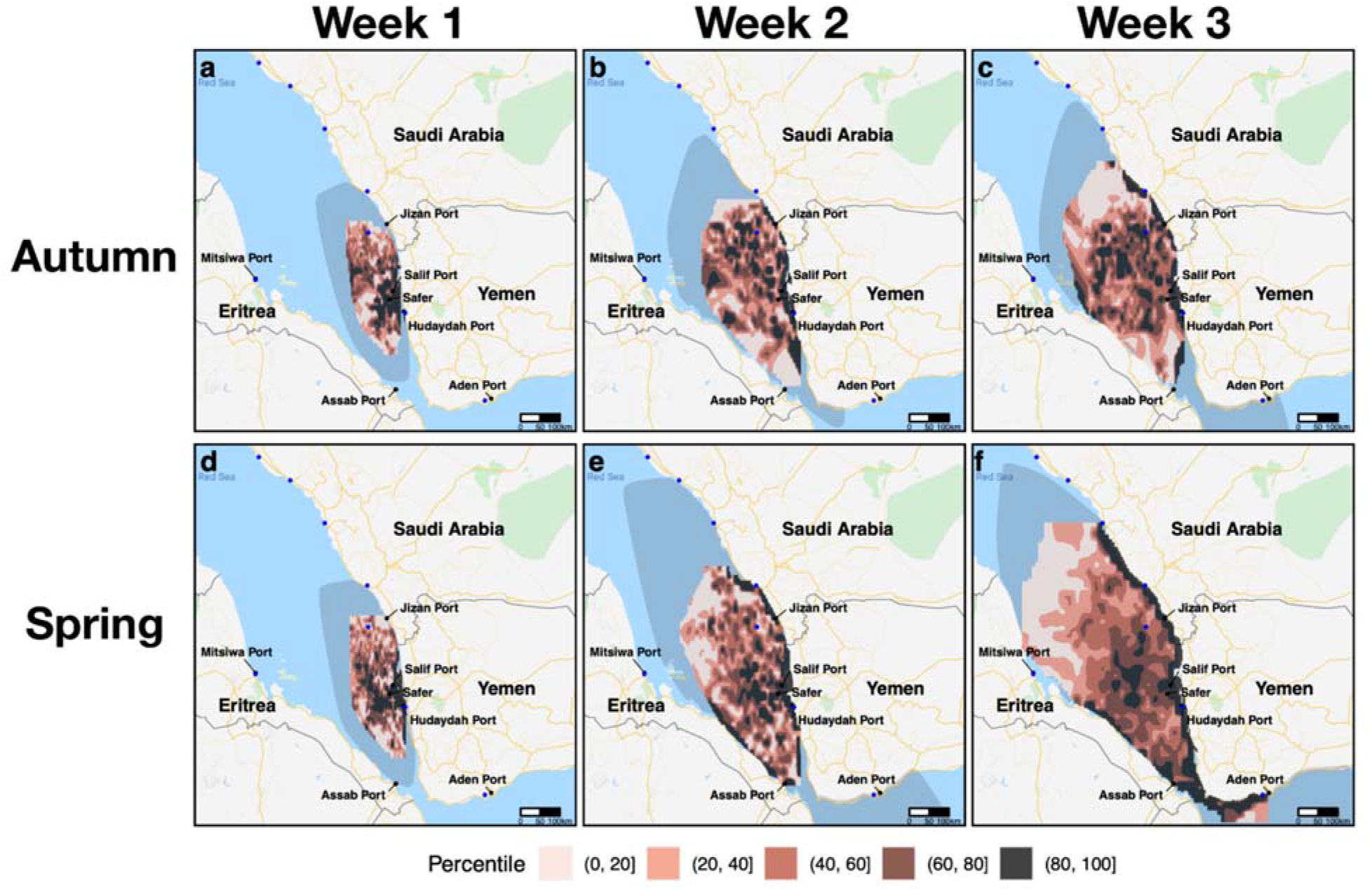
Average surface oil concentration of 1000 simulated spills in the autumn (a,b,c) and 1000 spills in the spring (d,e,f). Columns denote progress of the 1000 spills after one week (a,d), two weeks (b,e), and three weeks (c,f). Colored contours represent percentiles of average surface concentration over 1000 simulated spills. Shaded region represents the area within which approximately 90% of spill trajectories are expected to fall. Blue dots represent desalination plants.

**Supplementary Figure 3:**
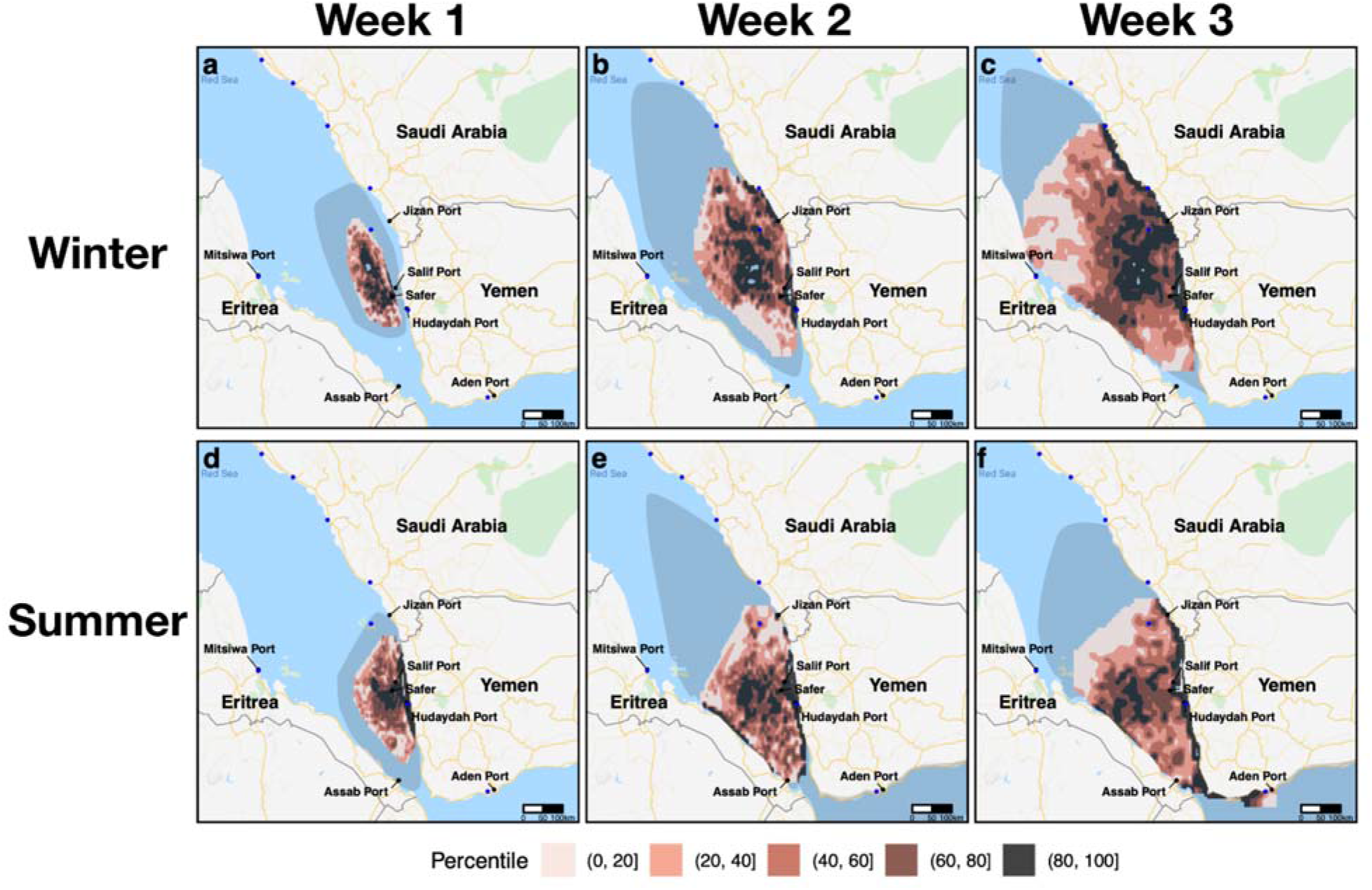
Average surface oil concentration of 1000 simulated spills in the winter (a,b,c) and 1000 spills in the summer (d,e,f), with the spatial resolution rounded to two decimal points instead of three. Columns denote progress of the 1000 spills after one week (a,d), two weeks (b,e), and three weeks (c,f). Colored contours represent percentiles of average surface concentration over 1000 simulated spills. Shaded region represents the area within which approximately 90% of spill trajectories are expected to fall. Blue dots represent desalination plants.

**Supplementary Figure 4:**
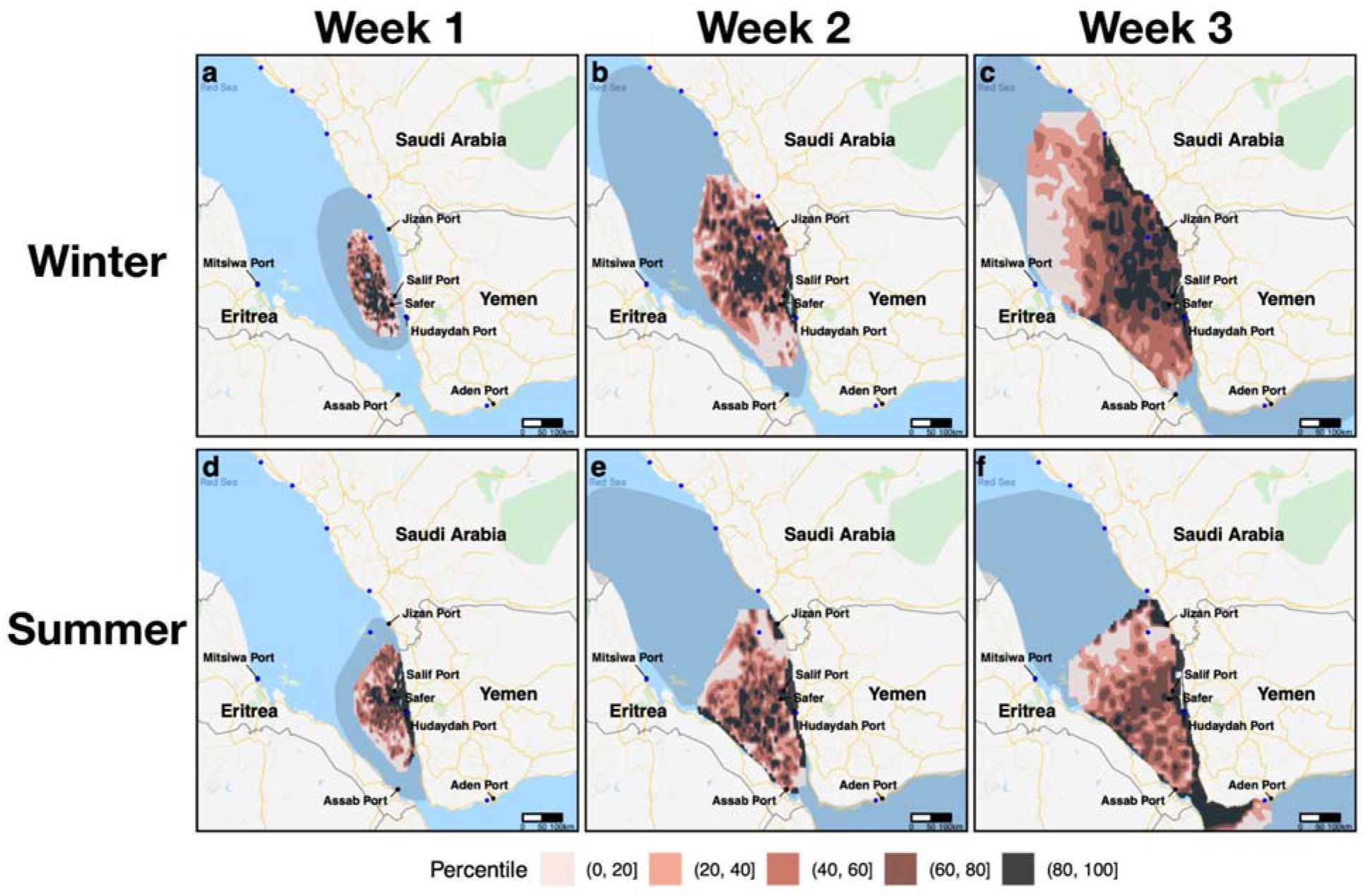
Average surface oil concentration of 1000 simulated spills in the winter (a,b,c) and 1000 spills in the summer (d,e,f), with 10,000 particles per spill instead of 1000. Columns denote progress of the 1000 spills after one week (a,d), two weeks (b,e), and three weeks (c,f). Colored contours represent percentiles of average surface concentration over 1000 simulated spills. Shaded region represents the area within which approximately 90% of spill trajectories are expected to fall. Blue dots represent desalination plants.

**Supplementary Figure 5:**
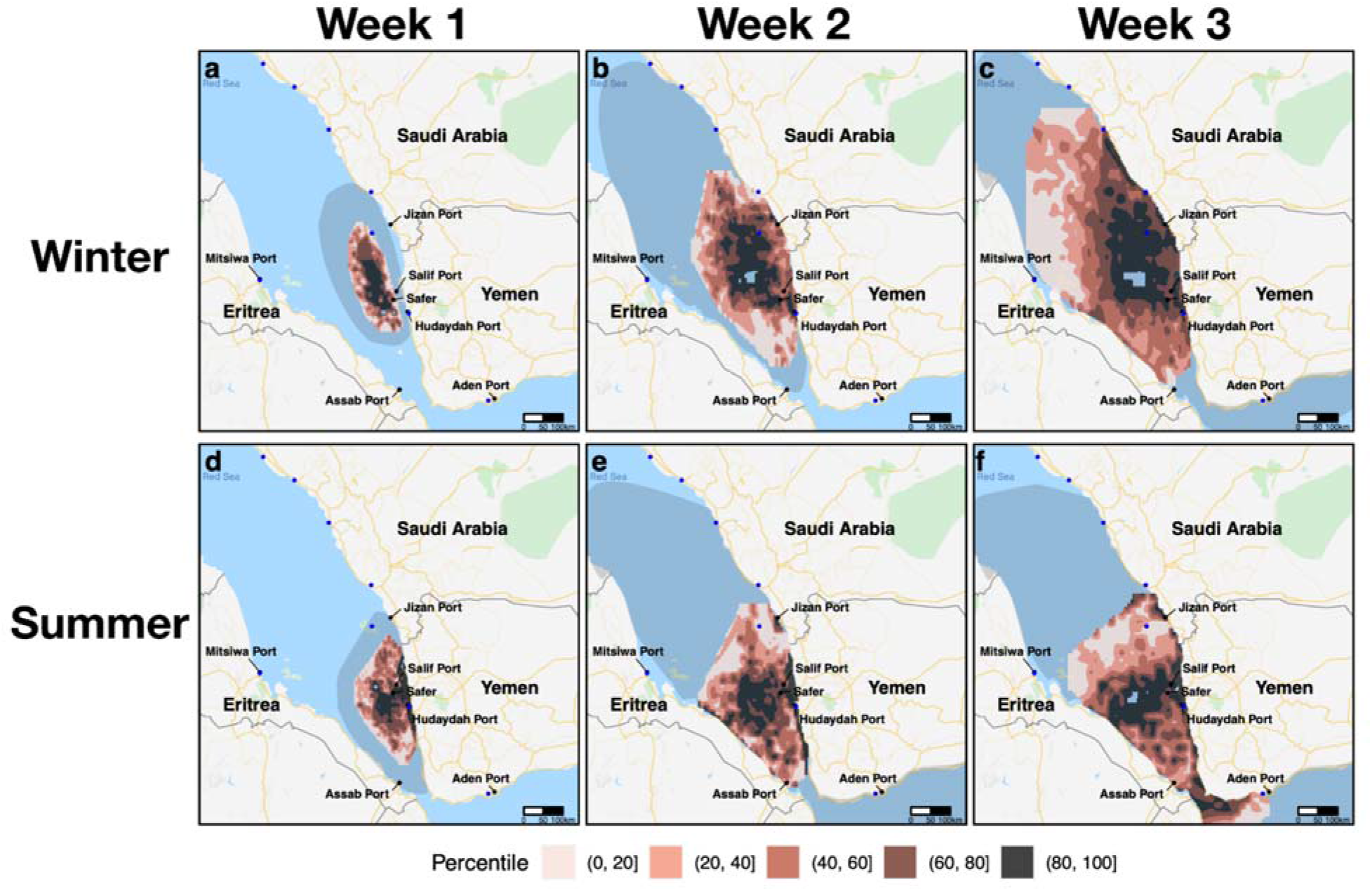
Average surface oil concentration of 1000 simulated spills in the winter (a,b,c) and 1000 spills in the summer (d,e,f), with the spatial resolution rounded to two decimal points and 10,000 particles per spill. Columns denote progress of the 1000 spills after one week (a,d), two weeks (b,e), and three weeks (c,f). Colored contours represent percentiles of average surface concentration over 1000 simulated spills. Shaded region represents the area within which approximately 90% of spill trajectories are expected to fall. Blue dots represent desalination plants.

**Supplementary Figure 6:**
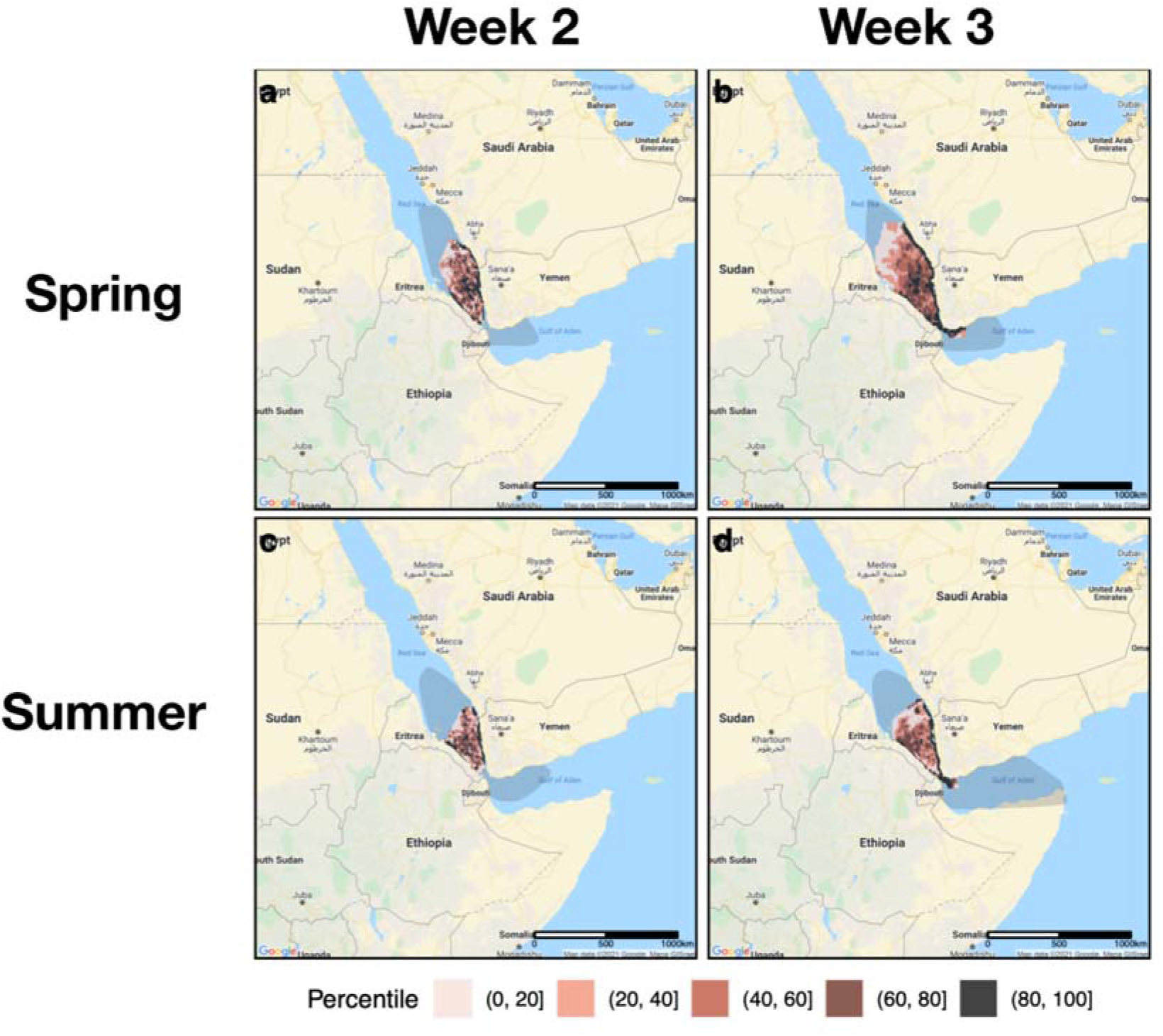
Zoomed out view of oil spill simulations for Spring (a,b) and Summer (c,d) in weeks 2 (a,c) and 3 (b,d). Colored contours represent percentiles of average surface concentration over 1000 simulations. Shaded region represents the area within which approximately 90% of spill trajectories are expected to fall, which is now visible for the Gulf of Aden.

**Supplementary Table 1:** Percentiles of oil surface concentration amongst exposed areas averaged over simulations at locations of interest.

| Location | Significance | Country | Week 1<br>Summer | Week 2<br>Summer | Week 3<br>Summer | Week 1<br>Winter | Week 2<br>Winter | Week 3<br>Winter |
| --- | --- | --- | --- | --- | --- | --- | --- | --- |
| Hudaydah | Port | Yemen | 94 | 96 | 94 | 0 | 98 | 97 |
| Salif | Port | Yemen | 84 | 90 | 88 | 0 | 91 | 94 |
| Aden | Port | Yemen | 0 | 0 | 26 | 0 | 0 | 0 |
| Hudaydah | Desalination | Yemen | 94 | 96 | 94 | 0 | 98 | 97 |
| Aden | Desalination | Yemen | 0 | 0 | 26 | 0 | 0 | 0 |
| Assab | Port | Eritrea | 0 | 23 | 24 | 0 | 0 | 0 |
| Jizan | Port | Saudi Arabia | 0 | 83 | 87 | 0 | 95 | 99 |
| Farasan | Desalination | Saudi Arabia | 0 | 55 | 66 | 0 | 65 | 70 |
| Shuqaiq | Desalination | Saudi Arabia | 0 | 0 | 0 | 0 | 0 | 95 |
| Qunfudah | Desalination | Saudi Arabia | 0 | 0 | 0 | 0 | 0 | 85 |

**Supplementary Table 2:** Locations and capacities of known desalination plants in the Southern Red Sea.

| Desalination plant | Country | Coordinates | Capacity (m<sup>3</sup>/day) |
| --- | --- | --- | --- |
| Hirgigo | Eritrea | 15.580140, 39.449509 | 2,000 |
| Farasan | Saudi Arabia | 16.687464, 42.096350 | 9,000 |
| Shuqaiq 1 | Saudi Arabia | 17.6599, 42.0761 | 94,000 |
| Shuqaiq 2 | Saudi Arabia | 17.6599, 42.0761 | 216,000 |
| Shuaiba | Saudi Arabia | 20.675714, 39.528004 | 1,707,540 |
| Qunfudah | Saudi Arabia | 19.125537, 41.078224 | 9,000 |
| Al Lith | Saudi Arabia | 20.130047, 40.266668 | 9,000 |
| Jeddah | Saudi Arabia | 21.480555, 39.177983 | 663,532 |
| Aden | Yemen | 12.745462, 44.836638 | 34,000 (estimated) |
| Hudaydah | Yemen | 14.800858, 42.966017 | 34,000 (estimated) |

**Supplementary Table 3:** Estimated percent of total fish yield loss for Yemen in the Red Sea region by week after spill and season of spill. Different levels of oil exposure, measured in terms of percentiles of surface concentration relative to other exposed areas, are selected as minimum values to cause fish yield loss.

|  | Week 1 | Week 2 | Week 3 |
| --- | --- | --- | --- |
| Winter, no threshold | 74.6 | 87.86 | 97.96 |
| Summer, no threshold | 90.02 | 100 | 100 |
| Winter, 10th percentile | 66.47 | 87.86 | 92.96 |
| Summer, 10th percentile | 85.2 | 97.96 | 100 |
| Winter, 20th percentile | 64.34 | 80.03 | 92.96 |
| Summer, 20th percentile | 77.62 | 95.47 | 100 |

**Supplementary table 4:**
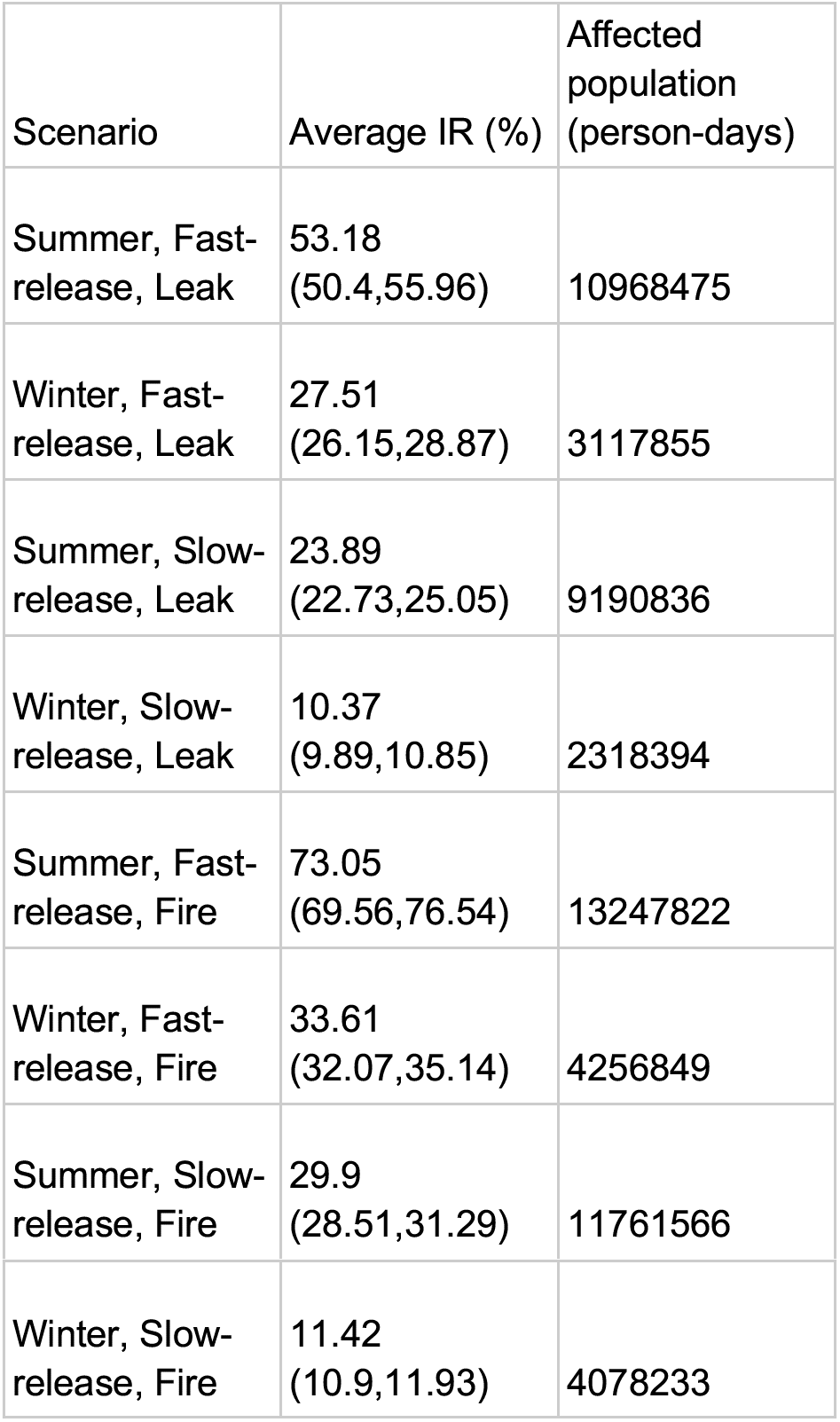
Population-weighted average increased risk and exposed populations for respiratory hospitalizations from air pollution over various scenarios and spill durations. Spill duration is equivalent to exposure duration. All intervals denote 95% confidence intervals. Exposed population is defined as having been exposed to 10 ug/m3 or more of PM2.5. Uses estimates from Wei et al.^1^

| Scenario | Average IR (%) | Affected population (person-days) |
| --- | --- | --- |
| Summer, Fast-release, Leak | 53.18<br>(50.4,55.96) | 10968475 |
| Winter, Fast-release, Leak | 27.51<br>(26.15,28.87) | 3117855 |
| Summer, Slow-release, Leak | 23.89<br>(22.73,25.05) | 9190836 |
| Winter, Slow-release, Leak | 10.37<br>(9.89,10.85) | 2318394 |
| Summer, Fast-release, Fire | 73.05<br>(69.56,76.54) | 13247822 |
| Winter, Fast-release, Fire | 33.61<br>(32.07,35.14) | 4256849 |
| Summer, Slow-release, Fire | 29.9<br>(28.51,31.29) | 11761566 |
| Winter, Slow-release, Fire | 11.42<br>(10.9,11.93) | 4078233 |

**Supplementary table 5:** Population-weighted average increased risk and exposed populations for cardiovascular and respiratory mortality from air pollution over various scenarios and spill durations. Spill duration is equivalent to exposure duration. All intervals denote 95% confidence intervals. Exposed population is defined as having been exposed to 10 ug/m3 or more of PM2.5. Uses estimates from Kloog et al.^2^

| Scenario | Average IR (%) | Affected population (person-days) |
| --- | --- | --- |
| Summer, Fast-release, Leak | 26.73<br>(25.93,27.53) | 10845335 |
| Winter, Fast-release, Leak | 13.55<br>(13.22,13.89) | 3163042 |
| Summer, Slow-release, Leak | 11.75<br>(11.42,12.07) | 9347517 |
| Winter, Slow-release, Leak | 5.04 (4.92,5.15) | 2272926 |
| Summer, Fast-release, Fire | 34.98<br>(34.11,35.85) | 13106374 |
| Winter, Fast-release, Fire | 16.68<br>(16.33,17.04) | 4360401 |
| Summer, Slow-release, Fire | 15.27<br>(14.91,15.63) | 11792403 |
| Winter, Slow-release, Fire | 5.64 (5.53,5.75) | 4066740 |

**Supplementary Table 6:** Quantities reported in the main text in their rounded form and originally calculated form.

| Quantity of interest | Reported estimate | Point estimate | 95% UI (Reported) | 95% UI (Calculated) |
| --- | --- | --- | --- | --- |
| Oil evaporated after 24 hours | 51 | 50.6 | 46-54 | 46.0275-53.9825 |
| Monthly fuel disruption | 200,000 | 206,723 | 180,000-250,000 | 184718.7-254009.7 |
| Monthly food aid disruption (Hudaydah only) | 5,700,000 | 5,684,800 | 3,700,000-8,100,000 | 3,695,800-8,122,600 |
| Monthly food aid disruption (Aden included) | 8,400,000 | 8,360,000 | 5,400,000-11,900,000 | 5,435,000-11,945,000 |
| Increased risk of cardiovascular<br>hospitalization at 1600ug/m3 | 530% | 529% | 460-590 | 464.2608-590.1955 |

